# Variation in uptake and dose reduction of CDK4/6 inhibitors for the treatment of breast cancer in England, 2019–2024: a descriptive observational study using OpenPrescribing Hospitals

**DOI:** 10.64898/2026.08.05.26359678

**Authors:** Louis Fisher, Calum Polwart, Christopher Wood, Ben Goldacre, Laura Anderson, Jennifer Isherwood, Sumeet Hindocha, Brian MacKenna, Victoria Speed

## Abstract

**Background:** The number of novel cancer therapies approved for use in England by the National Institute for Health and Care Excellence is increasing. Monitoring the adoption of new therapies is important to assess equity of access and evaluate real-world prescribing practices. OpenPrescribing Hospitals has recently been launched to facilitate analysis of open secondary care medicines data in England. Using this platform, we set out to describe the use of cyclin-dependent kinase 4 and 6 (CDK4/6) inhibitors, including the frequency of dose reductions, within National Health Service (NHS) hospitals in England between January 2019 and December 2024.

**Methods:** The monthly proportion of each CDK4/6 inhibitor relative to total CDK4/6 inhibitor use was calculated at hospital level. Regional variation was assessed across Cancer Alliances by comparing the proportions of each CDK4/6 inhibitor used within each alliance in 2021 and 2024. Use of lower strength palbociclib and abemaciclib was used as a proxy for dose reductions.

**Findings:** There was more than a 3-fold increase in the use of CDK4/6 inhibitors between 2019 and 2024. In 2019, 78.6%, 11.9% and 9.5% of CDK4/6 inhibitors used were palbociclib, abemaciclib and ribociclib, compared with 40.2%, 41.2% and 18.6% in 2024. There was variation in the relative percentage change in use of each agent by Cancer Alliance. Use of lower strengths was common for both palbociclib (60%) and abemaciclib (63%).

**Interpretation:** Changes in usage appeared responsive to publication of key evidence and regulatory milestones. There was a higher apparent frequency of dose reductions than reported in clinical trials. OpenPrescribing Hospitals is an accessible, publicly available tool for understanding uptake and use of medicines in NHS hospitals in England.

## Background

There is an increasing demand for Systemic Anti-Cancer Therapies (SACT), with the number of doses of SACT delivered in England increasing at 6-8% per year^1^. This is accompanied by significant innovation in available therapies. The average annual number of cancer drugs approved by the National Institute for Health and Care Excellence (NICE) has increased from 4.6 between 2000-2004 to 40.4 between 2020-2024^2^. This expansion reflects substantial progress in treatment options. Monitoring how these novel therapies are adopted into routine care in the NHS helps to ensure equitable access^3,4^ and allows evaluation of real-world prescribing practices, such as the frequency of dose reductions due to adverse reactions^5,6^, in comparison with controlled trial settings.

Breast cancer exemplifies the increasing availability of new therapies. Between 2002 and 2019, thirteen NICE technology appraisals for invasive disease received positive recommendations for use in England^7^. By the end of 2024, there were a further thirteen positive recommendations (**Table S1)**. Among these advances were the cyclin-dependent kinase 4 and 6 (CDK4/6) inhibitors: palbociclib, abemaciclib, and ribociclib. Following their market authorisation for the treatment of HR-positive, HER2-negative, locally-advanced or metastatic breast cancer by the European Medicines Agency between 2016-2018^8–10^, they were positively appraised by NICE for use within the NHS in England in the metastatic setting between 2017-2019^11–13^. Market authorisations and appraisals for use in early breast cancer came later and varied by agent: adjuvant abemaciclib was positively appraised for HR-positive, HER2-negative, lymph-node-positive disease in July 2022^14^; adjuvant treatment with ribociclib plus an aromatase inhibitor was positively appraised in July 2025 for HR-positive, HER2-negative early breast cancer at high risk of recurrence^15^; palbociclib remains unappraised for early breast cancer. These treatments were approved for the same metastatic indications within a short time period, and have diverging clinical evidence of effectiveness since their approval. A summary of the licensing, appraisals and landmark studies for progression free and overall survival (OS) is provided in **Box 1** and **Table S2**. For the majority of the period in which all three agents have been approved for use, there have been no national or international clinical guidelines recommending one agent over another for the same indication^16,17^. However, more recently, some guidance has begun to indicate agent preference in early^18^ and metastatic disease^19,20^. Given the complex timeline of licensing and appraisal, the evolving evidence base, and the emergence of guidance on agent preference, monitoring equitable access to these treatments is particularly important.

There are a range of tools for evaluating how medicines are adopted and used in routine NHS care in England. This includes: the Innovation Scorecard^21^, a bi-annual publication reporting use of medicines that have been positively appraised for use by NICE; the SACT Time to First Treatment Dashboard^22^, which reports the time taken for patients to be treated with SACT drugs in NHS trusts following NICE approval; and others^23–25^. We have extended this toolset by developing a publicly available platform using the openly available Secondary Care Medicines Dataset (SCMD)^26,27^, published monthly by the NHS. This supports transparent, timely and flexible analysis of medicines usage within the NHS in England. Here, we use the OpenPrescribing Hospitals platform to examine variation in uptake of CDK4/6 inhibitors in England, and to compare frequency of dose reductions between clinical trial settings and routine care. Specifically, we aimed to (i) examine the uptake of CDK4/6 inhibitors and variation in their use across NHS trusts (NHS organisations that provide and manage secondary healthcare services, which may include one or more hospital sites) and Cancer Alliances (groups of NHS organisations that work together to coordinate cancer care locally), and (ii) quantify the proportion of palbociclib and abemaciclib used at reduced doses in routine NHS practice.

#### Box 1. Timeline of NICE Technology Appraisals and landmark studies of clinical efficacy for palbociclib, abemaciclib, and ribociclib between 2016 and 2024.

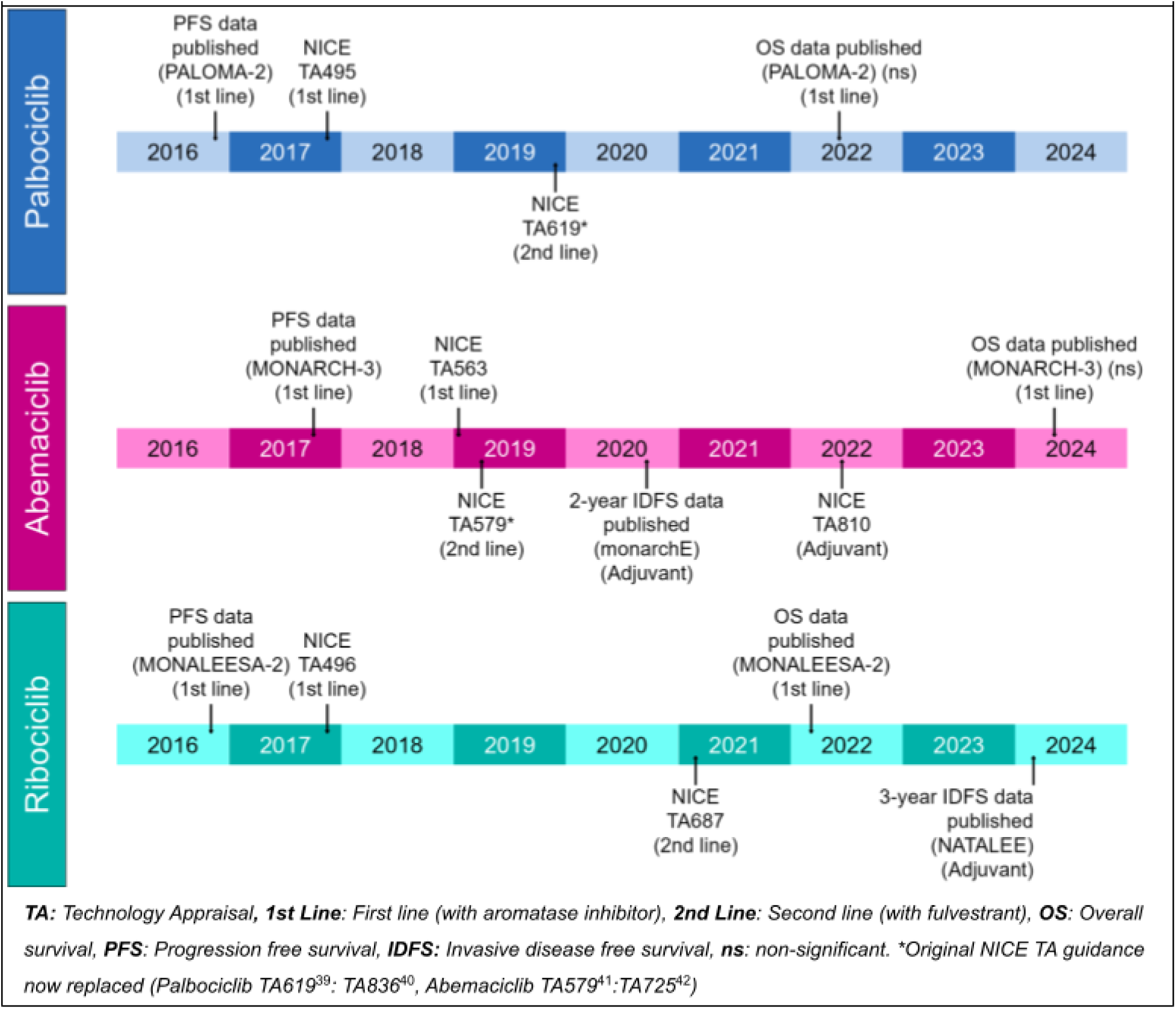

## Methods

We used the OpenPrescribing Hospitals platform to analyse the quantity of palbociclib, abemaciclib and ribociclib used within NHS trusts in England between January 2019 and December 2024^28,29^. CDK4/6 inhibitors were identified using the following Virtual Therapeutic Moiety (VTM) codes: 774399006 (*abemaciclib),* 777047009 (palbociclib) and 777435007 (ribociclib). The data available through the OpenPrescribing Hospitals platform is a curated aggregation of multiple data sources^26^. The primary data source is pharmacy stock control data from the SCMD^27^ that represents issuing of medicines to wards or clinical areas, which we refer to here as “use”. This data is additionally linked to: medicinal product information from the dictionary of medicines and devices (dm+d)^30^; trust information from the Organisational Data Service^31^; and medication classification information from the Anatomical Therapeutic Chemical/Defined Daily Dose (ATC/DDD) Index published by the World Health Organisation (WHO)^32^.

Individual trusts within the data are identified using Organisational Data Service codes^31^. Trusts are grouped by type (e.g. acute, mental health & learning disability, community) and the Cancer Alliance they are affiliated to^33–35^. Only trusts with any CDK4/6 inhibitor use between January 2019 and December 2024 are included. Mental health trusts, community trusts, and trusts not partnered with a Cancer Alliance were excluded from the analysis. We report the monthly usage of each CDK4/6 inhibitor product using Defined Daily Doses (DDDs). A DDD is defined by the WHO as the assumed average maintenance dose per day for a drug used for its main indication in adults, providing a unit of drug utilisation that allows comparison across preparations^36^. To assess trust-level variation in CDK4/6 inhibitor use, we calculate the monthly proportion of each CDK4/6 inhibitor as a share of total CDK4/6 inhibitor use within each trust. Each month, trusts are ranked and percentiles are calculated. These percentiles are presented as time series charts. Where appropriate, charts are annotated with the date of NICE appraisal decision and dates of additional evidence publication. To assess variation by Cancer Alliance we calculated the percentage of each CDK4/6 inhibitor as a proportion of all CDK4/6 inhibitors across all trusts within each Cancer Alliance in 2021 and 2024, and assessed the change across this period. 2021 was chosen as a suitable comparator period in which all three agents had had NICE approval with an aromatase inhibitor for previously untreated, hormone receptor-positive, HER2-negative, locally advanced or metastatic breast cancer. Later in 2021, ribociclib (March 2021) and then abemaciclib (September 2021) were approved for treating hormone receptor-positive, HER2-negative advanced breast cancer after endocrine therapy.

The licensed dosing schedule for each CDK4/6 inhibitor is described in more detail in **Table S3**. For abemaciclib and palbociclib, we calculated the proportion of reduced strength preparations as a proxy for the frequency of dose reductions between January 2024 and December 2024. We are not able to determine dose reductions for ribociclib as only a single strength of tablet was available during the study period.

## Results

We identified ten CDK4/6 inhibitor Virtual Medicinal Products (VMPs) reported in the SCMD (**Table 1**) used by 109 eligible NHS trusts between January 2019 and December 2024 **(Figure S1)**. The included and excluded trusts are shown in **Table S4** and **Table S5.** The total DDDs used for each VMP in 2019, 2024, and January 2019-December 2024 are shown in **Table 1**.

**Table 1.**
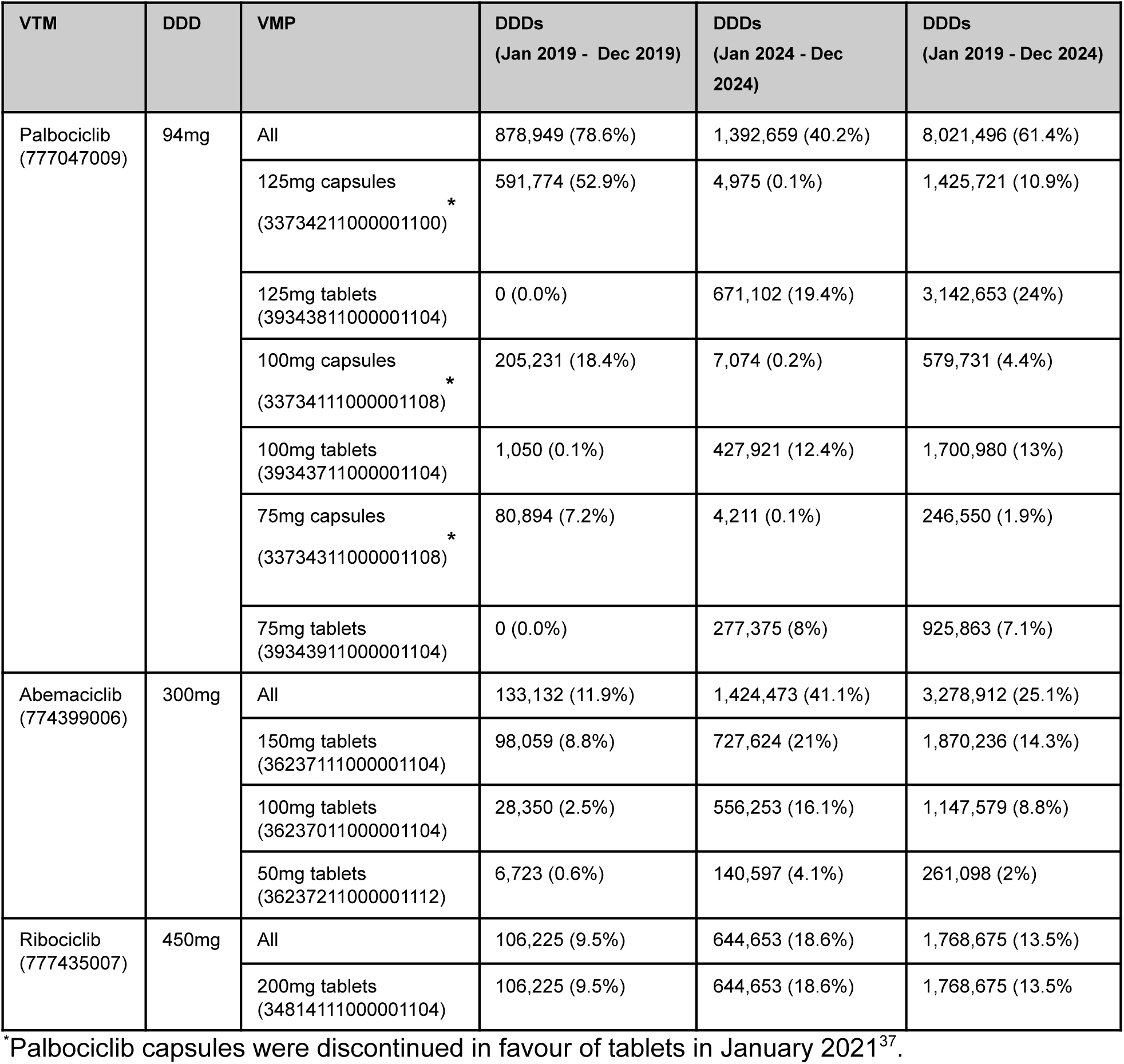
Summary of total Defined Daily Doses (DDDs) used by Virtual Therapeutic Moiety (VTM) and Virtual Medicinal Product (VMP) between: January 2019-December 2019, January 2024-December 2024 and January 2019-December 2024.

| VTM | DDD | VMP | DDDs<br>(Jan 2019 - Dec 2019) | DDDs<br>(Jan 2024 - Dec 2024) | DDDs<br>(Jan 2019 - Dec 2024) |
| --- | --- | --- | --- | --- | --- |
| Palbociclib<br>(777047009) | 94mg | All | 878,949 (78.6%) | 1,392,659 (40.2%) | 8,021,496 (61.4%) |
|  |  | 125mg capsules<br>(33734211000001100) * | 591,774 (52.9%) | 4,975 (0.1%) | 1,425,721 (10.9%) |
|  |  | 125mg tablets<br>(39343811000001104) | 0 (0.0%) | 671,102 (19.4%) | 3,142,653 (24%) |
|  |  | 100mg capsules<br>(33734111000001108) * | 205,231 (18.4%) | 7,074 (0.2%) | 579,731 (4.4%) |
|  |  | 100mg tablets<br>(39343711000001104) | 1,050 (0.1%) | 427,921 (12.4%) | 1,700,980 (13%) |
|  |  | 75mg capsules<br>(33734311000001108) * | 80,894 (7.2%) | 4,211 (0.1%) | 246,550 (1.9%) |
|  |  | 75mg tablets<br>(39343911000001104) | 0 (0.0%) | 277,375 (8%) | 925,863 (7.1%) |
| Abemaciclib<br>(774399006) | 300mg | All | 133,132 (11.9%) | 1,424,473 (41.1%) | 3,278,912 (25.1%) |
|  |  | 150mg tablets<br>(36237111000001104) | 98,059 (8.8%) | 727,624 (21%) | 1,870,236 (14.3%) |
|  |  | 100mg tablets<br>(36237011000001104) | 28,350 (2.5%) | 556,253 (16.1%) | 1,147,579 (8.8%) |
|  |  | 50mg tablets<br>(36237211000001112) | 6,723 (0.6%) | 140,597 (4.1%) | 261,098 (2%) |
| Ribociclib<br>(777435007) | 450mg | All | 106,225 (9.5%) | 644,653 (18.6%) | 1,768,675 (13.5%) |
|  |  | 200mg tablets<br>(34814111000001104) | 106,225 (9.5%) | 644,653 (18.6%) | 1,768,675 (13.5%) |

\*Palbociclib capsules were discontinued in favour of tablets in January 2021<sup>37</sup>.

Between January 2019 and December 2024, there were a total of 13,069,083 DDDs of CDK4/6 inhibitors used. Palbociclib was the most frequently used across this period, accounting for 8,021,496 DDDs (61.4%).

There was an increasing trend in the total volume of CDK4/6 inhibitors used between January 2019 and December 2024 (**Figure 2, Table S6**). In 2019, palbociclib was the most frequently and most commonly used CDK4/6 inhibitor. Following the adjuvant approval of abemaciclib for the treatment of early breast cancer by NICE in July 2022^14^, there has been a consistent increasing trend in its usage and as of December 2024 it showed similar volume of usage as palbociclib (abemaciclib DDDs in 2024 1,424,473 vs palbociclib DDDs in 2024 1,392,659). Following publication of positive OS data for ribociclib in patients with HR-positive, HER2-negative advanced breast cancer in March 2022 (MONALEESA-2)^38^, and subsequent no OS benefit data for palbociclib in June 2022 (PALOMA-2)^39^, an increase in ribociclib and a decrease in palbociclib use was observed (**Figure 2**).

**Figure 2.**
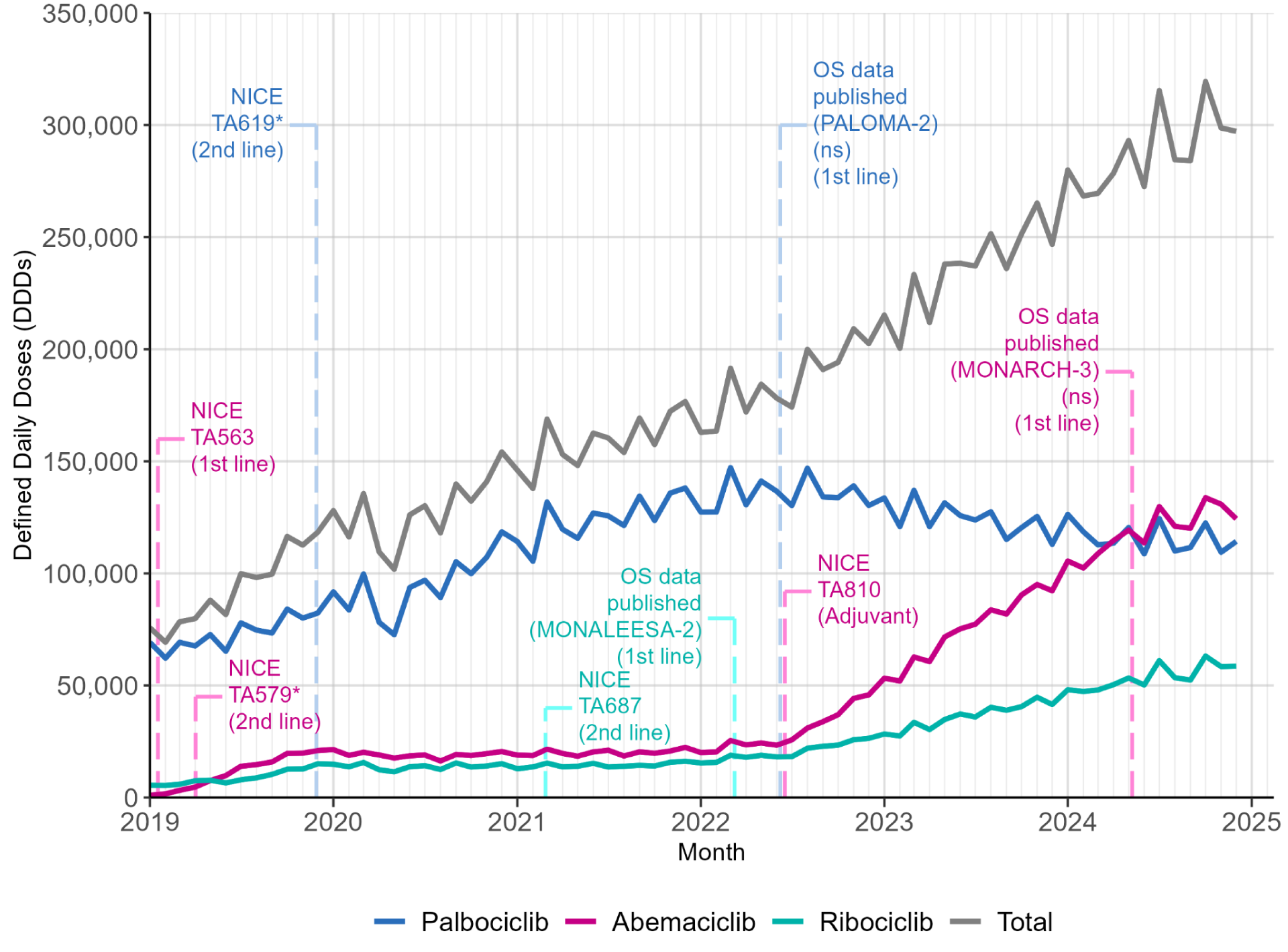
Monthly Defined Daily Doses (DDDs) of abemaciclib, palbociclib and ribociclib used across all eligible NHS trusts in England between January 2019 and December 2024. Key dates related to changes in the evidence base or approvals for these medications are highlighted. *\*Original NICE TA guidance, now replaced (Palbociclib TA619*^40^*: TA836*^41^*, Abemaciclib TA579*^42^*:TA725*^43^)

### Trust-level variation

The trust level variation in the proportion of each CDK4/6 inhibitor used is shown in **Figure 3**. The median proportion of palbociclib used across the study period decreased from 100% in January 2019 to 38% in December 2024 (**Figure 3a**). In the same period, the median trust-level proportion of abemaciclib **(Figure 3b)** and ribociclib **(Figure 3c)** used increased from 0% to 40% and 19%. Whilst there is relatively consistent variation across this time period, there are differences in the rate of uptake. Example patterns for individual trusts are shown in **Figure 3**, highlighting varying magnitudes and speed of change in response to new appraisals and evolving evidence.

**Figure 3.**
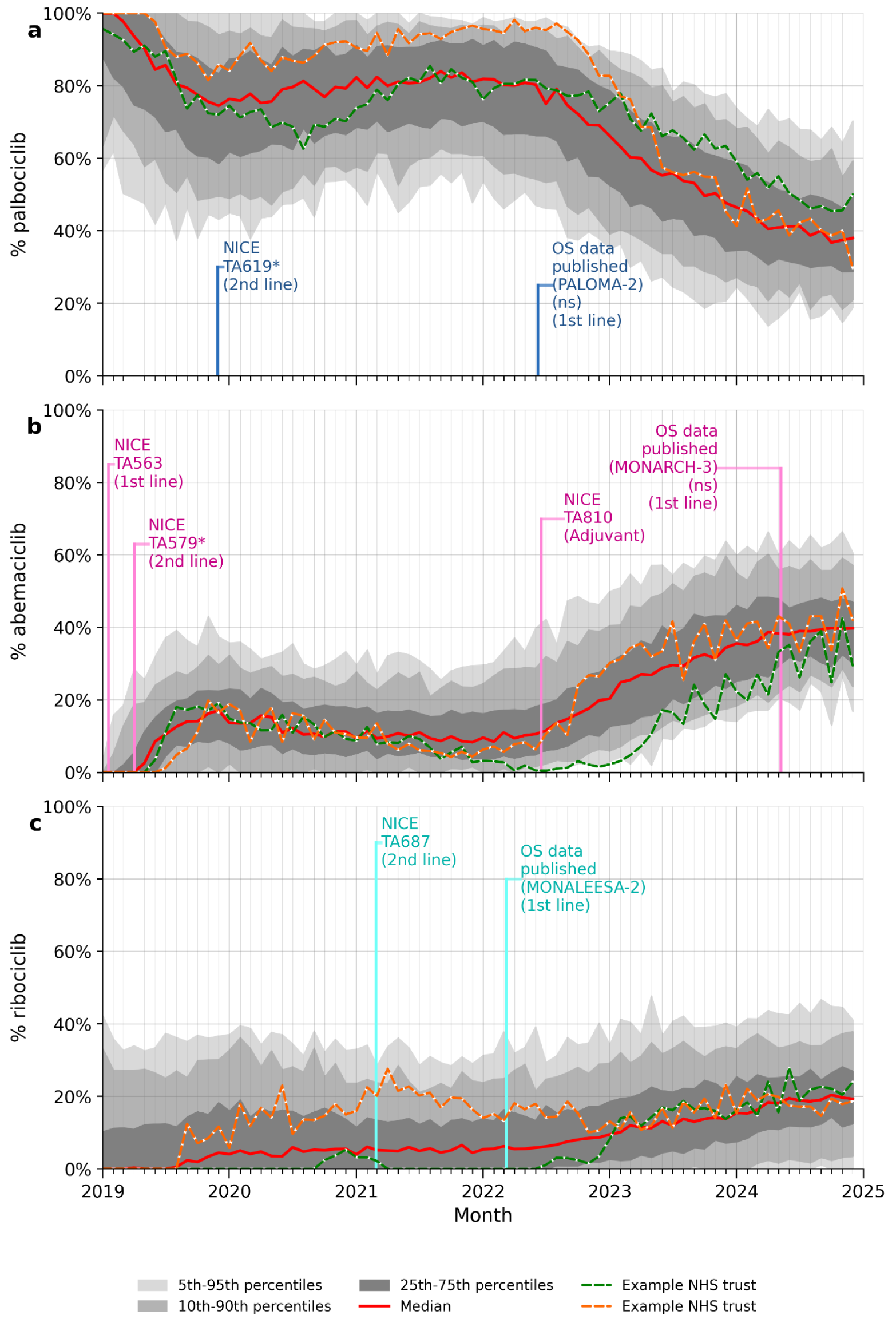
Monthly proportion of **a)** palbociclib **b)** abemaciclib **c)** ribociclib DDDs relative to total CDK4/6 inhibitor DDDs at NHS trust level between January 2019 and December 2024 . Grey bands show quantiles (25-75%, 10-90% and 5-95%) calculated across all eligible NHS trusts. The solid red line is the median. Example individual trust issuing patterns are shown by orange and green dotted lines. *\*Original NICE TA guidance, now replaced (Palbociclib TA619*^40^*: TA836*^41^*, Abemaciclib TA579*^42^*:TA725*^43^)

### Variation by Cancer Alliance

The proportion of each CDK4/6 inhibitor used within each Cancer Alliance in 2021 and 2024, is shown in **Figure 4**. The percentage used as palbociclib in 2021 ranged from 65% in the South East London Cancer Alliance to 95% in the North East London Cancer Alliance (**Table S7**). The percentage of palbociclib used in 2024 was lower, ranging from 25% in the South East London Cancer Alliance to 55% in the Cheshire and Merseyside Cancer Alliance. The percentage point change in palbociclib used between 2021 and 2024 ranged from -15% in the Humber and North Yorkshire Cancer Alliance to -56% in the North Central London Cancer Alliance. The percentage point change in abemaciclib used in the same period ranged from +15% in the Humber and North Yorkshire Cancer Alliance to +44% in the Greater Manchester Cancer Alliance. This change may be accounted for by the introduction of adjuvant abemaciclib and expansion of the eligible population to include patients with early breast cancer, however, there has also been a transition to use of more ribociclib in most cancer alliances; the percentage point change in ribociclib used in this period ranged from -2% in the East Midlands Cancer Alliance to +21% in the the Northern Cancer Alliance.

**Figure 4.**
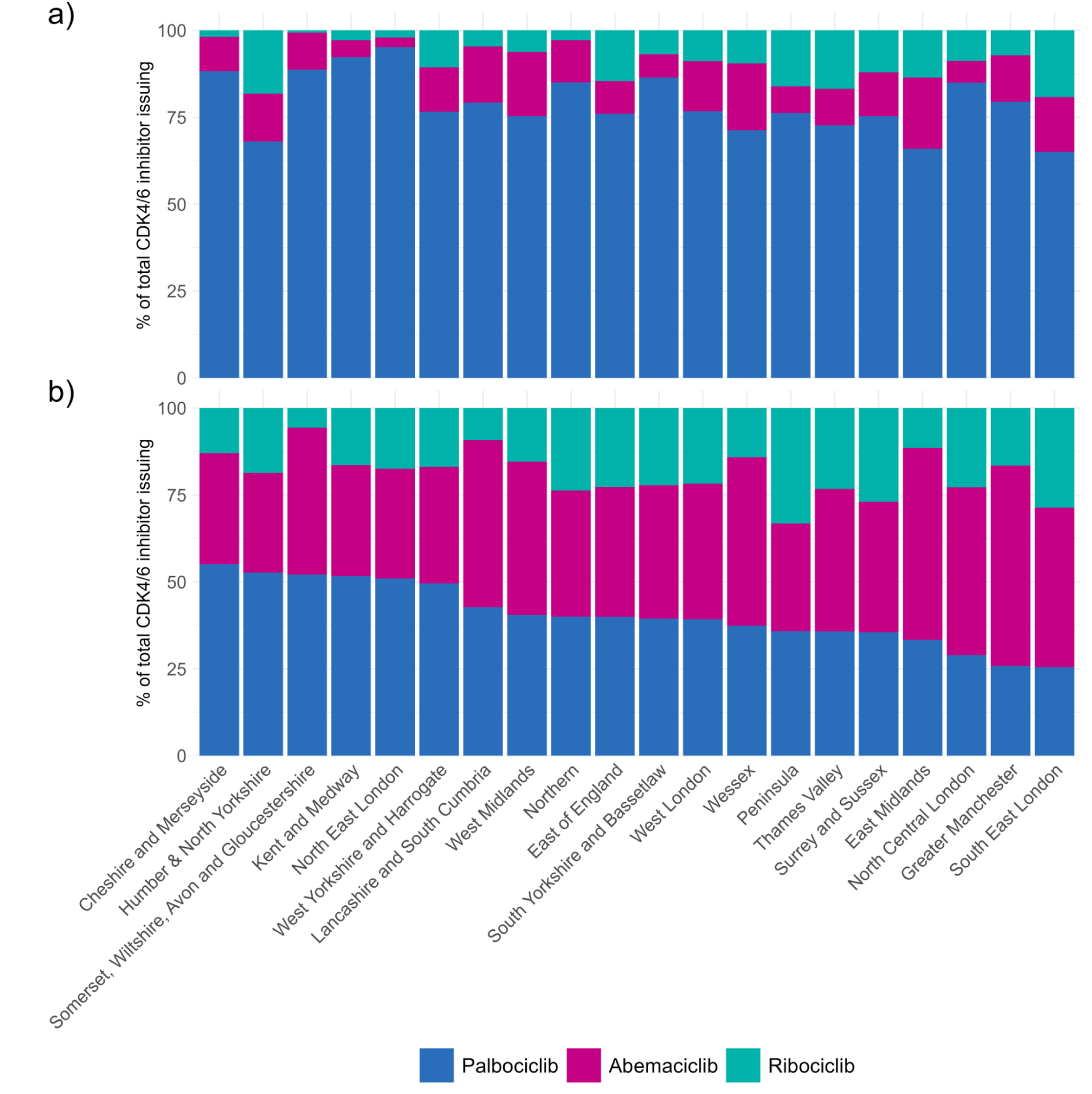
Proportion of each CDK4/6 inhibitor used across all eligible trusts in each NHS Cancer Alliance between **a)** January 2021 and December 2021 **b)** January 2024 and December 2024.

### Dose reduction data

**Figure 5** shows the proportion of tablets used of each strength for abemaciclib and palbociclib between January 2024 and December 2024. 63.3% of abemaciclib tablets used were low strength (100mg or 50mg). 60% of palbociclib tablets were low strength (100mg or 75mg).

**Figure 5.**
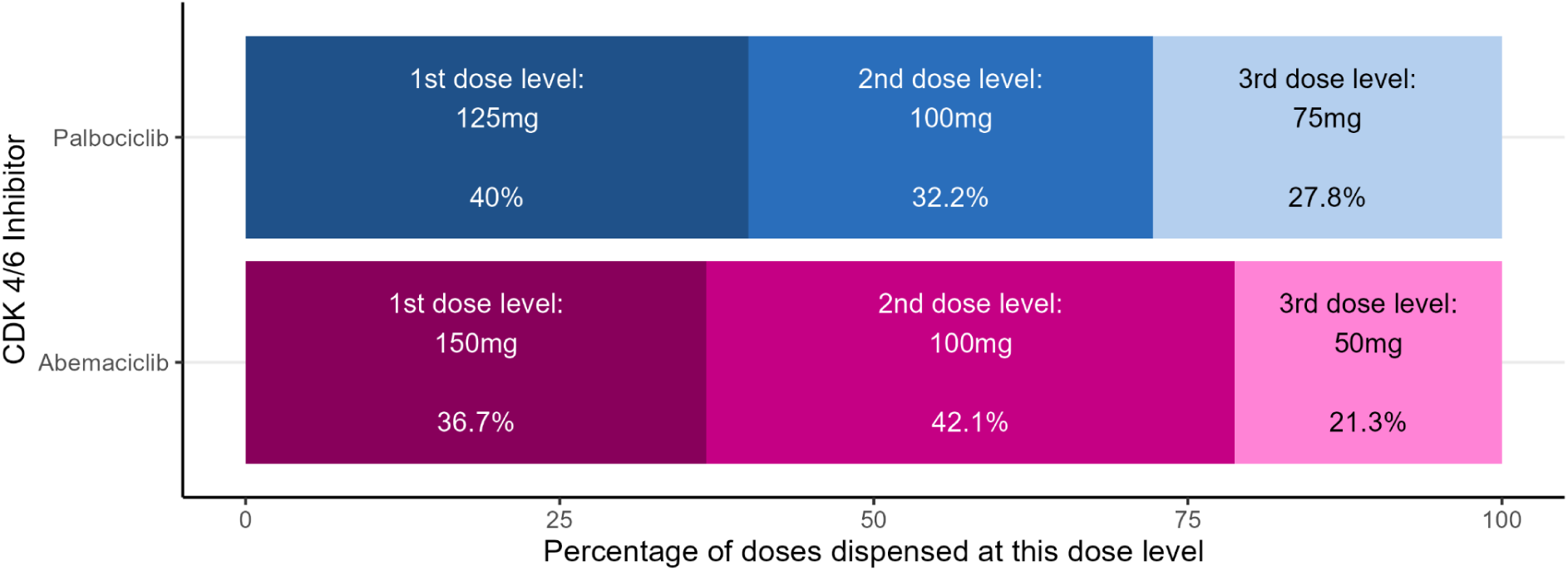
Proportion of abemaciclib and palbociclib tablets used across all trusts by strength between January 2024 and December 2024.

## Discussion

This study reports more than a 3-fold increase in the use of CDK4/6 inhibitors in England between 2019 and 2024, and shows changing patterns in agent choice. For example, palbociclib, which accounted for 78.6% of all CDK4/6 inhibitors used in 2019 (11.9% abemaciclib; 9.5% ribociclib), fell to 40.2% of all issuing in 2024 (41.1% abemaciclib, 18.6% ribociclib). This varied by Cancer Alliance. For example by 2024, the proportion of abemaciclib used in the Greater Manchester Cancer Alliance was twice as high as that of the Humber and North Yorkshire Cancer Alliance. Use of lower strengths was common for both palbociclib (60%) and abemaciclib (63%), suggesting a higher frequency of dose reductions than observed in phase III licensing studies.

Changes in CDK4/6 use appear responsive to changes in key evidence and regulatory approvals. At the start of the study period, palbociclib was the most widely used CDK4/6 inhibitor despite the simultaneous positive appraisal by NICE for ribociclib in November 2017. This likely reflects greater initial clinical familiarity with palbociclib as a result of its earlier international approval^44,45^ and UK early-access schemes^46^. Palbociclib use peaked in early 2022 and then declined around the time of non-significant OS results from PALOMA-2 in June 2022^39^. Ribociclib use increased more gradually throughout the study period, with a steeper increase starting at the time of publication of favourable OS data from MONALEESA-2 in March 2022^38^. The sharp rise in abemaciclib from mid-2022 coincides with the positive NICE appraisal for adjuvant use (TA810^14^), and may therefore partly reflect expansion of the eligible patient population rather than substitution of agents in the metastatic setting. Additional factors influencing agent choice may include patient preference, toxicity profiles and local implementation processes.

This study highlights the value of an open, easy-to-use tool for understanding real-world medicines usage: how they are used, how their use varies, and how they are used outside of a controlled clinical trial setting.

### Strengths and Weaknesses

This is the first study to examine national use of CDK4/6 inhibitors in England following the availability of post-marketing OS data for all three licensed agents. By reporting results between 2019 and 2024, the study captures a time period with numerous approvals, extensions to licensed indications and heterogeneous trial results. Additionally, we highlight variation in the CDK4/6 agent of choice at a range of organisational levels including NHS trusts, and Cancer Alliance. Examining variation at different levels may help determine whether the barriers and facilitators for adopting novel therapies are predominantly local or more widely shared. Another strength is the use of routinely collected pharmacy stock data with national coverage, analysed using an open platform with transparent and reproducible methods^47^. This work highlights the potential for the OpenPrescribing Hospitals platform to support organisations to undertake local audit and quality improvement activities without the need for additional data collection.

However, there are limitations in the use of aggregate pharmacy stock control data. First, we cannot determine whether these medicines have been used for indications outside of NICE recommendations. The NHS will only reimburse these medicines where they have been used in line with NHS England’s prior authorisation process, but there may be a small amount of other use, either privately funded or supplied through trials or pharmaceutical company access schemes^46^. Second, the completeness of the data within the SCMD is dependent on submissions by individual NHS trusts. Incomplete submissions may result in underestimation of CDK4/6 issuing in some NHS trusts^48^. Trusts are only included in this analysis if they have issued any of the identified CDK4/6 inhibitors. If a trust held stock, but did not issue the products during the study period, they were not included. Third, we were unable to differentiate if a CDK4/6 inhibitor was issued but not administered to a patient. Fourth, using the aggregate stock control data available in the SCMD it is not possible to determine if patients commence on a reduced dose or are subsequently dose reduced, however, there is no circumstance in the Summary of Product Characteristics for these products where a lower starting dose is recommended^49–51^.

Beyond limitations with the data used, secondary care organisations do not have strictly defined patient populations, limiting our ability to apply a meaningful population, disease, or activity-based denominator. This restricts interpretation of variation between trusts and Cancer Alliances; observed variation may also be reflective of differences in access, clinician choice, number of eligible patients, and the case-mix of eligible patients. Variation by Cancer Alliance should also be interpreted cautiously as trusts may provide care for patients living outside their local Cancer Alliance boundary. It is possible that some of the abemaciclib and ribociclib used prior to their approval for use in early breast cancer (in June 2022 and July 2025), represents trial use and that some variation by Cancer Alliance is explained by trial participation. Finally, the data in this study is most likely to represent use in the metastatic setting with the NICE approvals for early breast cancer published later in the study period.

### Findings in Context

For most of the study period, in the absence of national or international guidance recommending one CDK4/6 inhibitor over another, observed variation likely reflects local decision-making, implementation and monitoring capacity, and response to emerging evidence. At the time of writing, each agent is similarly priced in the NHS based on indicative cost^52^, although this may not reflect the cost paid by the NHS based on confidential price agreements^53^. Understanding other factors that can drive the observed variation can help ensure equitable access. These factors may reflect differences in organisational capacity and resources to implement practice changes, changing evidence, formulary status, familiarity with a particular agent, historical prescribing patterns, and differences in patient case mix with regards to toxicity profiles and tolerability.

In this context, the ability to identify which treatments are being used, where they are being used, and to benchmark their use across organisations is essential for monitoring adoption across the NHS in England. The results of this study build on an interrupted time series to investigate the impact of NICE technology appraisals for invasive breast cancer by Gannon et al^7^. The study linked data collected in the National Audit of Breast Cancer in Older Patients and SACT data in England^7^. Using pseudonymised cancer registration patient records for all women aged 50 years and over diagnosed with breast cancer between January 2014 and December 2019, this showed 87.2% of CDK4/6 inhibitor based first-line treatment was palbociclib, but there was insufficient uptake of ribociclib and abemaciclib to carry out further analyses. This aligns with our results for the same period. Gannon and colleagues suggest that SACT data returns may also be low for some NHS trusts and as such estimates for the use of new drugs for recently diagnosed patients may be higher. Data in the SCMD is submitted routinely by individual trusts for processed pharmacy stock, providing an alternative to SACT data returns.

Dose reductions for abemaciclib appear more common in our study than seen in the clinical trial setting. In MONARCH-2^54^ and -3^55^, the licensing trials for abemaciclib as a second-line and first-line treatment for metastatic breast cancer, the rate of dose reductions was 42.9% and 43.4%^56,57^. Similarly, 43.6%^58^ of patients were dose reduced in the MonarchE trial^59^, the licensing trial for abemaciclib as an adjuvant treatment for early breast cancer. In contrast, throughout 2024, we found that 63.3% of abemaciclib used in trusts in England were lower strength preparations (100mg or 50mg tablets). This is consistent with other real-world data. In a UK multi-centre study of first- or subsequent-line treatment for metastatic breast cancer, dose reductions were observed in 58.8% of patients receiving abemaciclib^60^. In England, a single centre study (n=40) found dose reductions in 62.5% of patients that received adjuvant abemaciclib^61^. A separate retrospective study in England reported dose reductions of 51% in 55 patients treated in the same context^62^.

As for abemaciclib, we observed a higher frequency of dose reductions for palbociclib (60%) than reported from the licensing studies. In the same multi-centre study of metastatic breast cancer treatment described above, a dose reduction rate of 53.8% was observed for patients receiving palbociclib^60^. The PALOMA-2^63^ and -3^64^ studies, the licensing trials for palbociclib as a second-line and first-line treatment for metastatic breast cancer, had dose reduction rates of 36% and 34%^65,66^. Other studies show results more comparable with those from trial settings: real-world usage patterns of palbociclib in the Netherlands between 2017 and 2020 showed a dose reduction rate of 33%^67^; a retrospective analysis of a US nationwide dataset found 40.2 % of patients receiving palbociclib had dose adjustments^68^.

Whilst these measures may not be directly comparable as we report stock issued rather than patient-level dose reductions, our results suggest a greater use of dose modification in routine care in the NHS in England. Dose reductions have been shown to be an effective strategy for management of adverse events while allowing continued exposure and optimising the therapeutic index^5,6,69,70^. Adverse event grading can be subjective and clinicians may apply different thresholds for reducing doses, particularly for toxicities such as diarrhoea. This is likely to result in a greater proportion of patients receiving a dose reduction while enabling patients to remain on treatment. The observed differences could also reflect differences in patient population.

### Policy Implications

Through publication of the SCMD, produced by Rx-Info^71^ and hosted by the NHS Business Services Authority^72^, the NHS is the first health service in the world to make data for the use of cancer medicines across an entire country publicly available. This study shows that this is a rich resource for understanding real-world patterns in medicines usage.

As the number of NICE-approved oncology therapies continues to increase, ensuring timely and equitable access across the NHS is a growing priority^73^. This requires appropriate, transparent benchmarking and comparison that allows individuals to quickly see how their own organisation compares with other trusts, and supports identification of differences in treatment adoption that may otherwise remain unrecognised. Smaller trusts, often operating with more constrained resources, may find learning from bigger cancer centres, who are often the most visible in the field through teaching and conference presentations, is not always realistic or transferable to their setting. The same challenge applies to trusts serving older populations or communities with high levels of comorbidity. Easy access to medicines usage data coupled with local knowledge gives autonomy to local organisations to act appropriately to improve care.

Whilst access to national data is welcome, there are further developments which could support improved clinical audit and evaluation of cancer care. Firstly, access to comparable open data from other countries would be valuable to understand how different health systems adopt novel therapies and could inform national and local decision-making and support shared learning. Secondly, the NHS collects detailed patient level information through its reimbursement system for managing approval of certain medicines, commonly called “High Cost Drugs”^74^. However, this information is not collated at a national level or made widely available internally or externally in the NHS for clinical audit and evaluation of cancer care. An aggregated NHS high-costs drugs dataset including indication for use and homecare status would augment the currently open data, benefitting this study and many others.

### Future Research

There is variation in the uptake of individual CDK4/6 inhibitors in response to the latest approvals and OS data. Further qualitative research is needed to understand what drives faster adoption of novel therapies. The OpenPrescribing Hospitals platform could be used to identify targets for interventions to speed up the adoption of new treatments. For example, by identifying those with rapid uptake and sharing best practice methods with those identified as having slower uptake^75^.

The observed shifts in dominant agent largely predate formal guidelines recommending one agent over another. There has since been updated guidance published by the National Comprehensive Cancer Network (June 2023^19^) and European Society for Medical Oncology (May 2026^20^) in the metastatic setting and American Society of Clinical Oncology (May 2024^18^) in the early breast cancer setting. Continued geographic variation in agent choice emphasises the need for ongoing monitoring as guidance evolves.

Ribociclib was approved with an aromatase inhibitor for adjuvant treatment of hormone receptor-positive HER2-negative early breast cancer at high risk of recurrence in April 2025 (TA1086)^15^. Following initial approval restricted to patients with lymph node-positive disease, the eligible population was later expanded to include patients with no nodal involvement but with other high risk features including high genomic risk or high Ki67. Continued monitoring of the patterns of CDK4/6 inhibitor use in England could help limit inequities caused by known barriers to accessing genomic testing^76,77^.

## Conclusion

Using the OpenPrescribing Hospitals platform, we identified: variation in uptake of CDK4/6 inhibitors between Cancer Alliances; changes in their usage around the time of publication of key evidence and regulatory milestones; and a higher apparent frequency of dose reductions than reported in clinical trials. These findings highlight the value of openly available secondary care medicines data and demonstrates how the OpenPrescribing Hospitals platform can be used to provide timely, national insight into the adoption of newly approved cancer therapies in the NHS in England.

## Supporting information

RECORD statement

## Data Availability

The underlying data used for this for this analysis is provided by Rx-Info and published under an Open Government Licence by the NHS Business Services Authority. It is accessed through the openly available OpenPrescribing Hospitals platform. Intermediate data, and the code for data management and analysis, are openly available for inspection and re-use at github.com/ebmdatalab/cdk4-6-uptake-openprescribing-hospitals under an MIT licence.

## Ethical approval

No ethical approval was sought for this analysis as this study uses open, publicly available, and anonymised data.

## Patient and Public Involvement

OpenPrescribing Hospitals is an openly accessible data explorer for secondary care medicines data, which frequently receives user feedback from professionals, patients and the public. This feedback is used to refine and prioritise our informatics tools and research activities. Patients were not formally involved in developing this specific study design

## Guarantor

VS is guarantor

## Contributorship

**Conceptualization**: CP, LF

**Data curation**: LF,CP

**Formal analysis**: CP,LF

**Funding acquisition**: BMK,BG

**Investigation**: CP,LF

**Methodology**: LF,CP,CW,VS

**Project administration**: VS

**Resources**: BMK, BG

**Software**: CP,LF

**Validation**: LF,CP,VS

**Visualization**: CP,LF, VS

**Writing - original draft**: CP,LF,VS,LA,JI,SH

**Writing - review & editing**: CP,LF,CW,LA,JI,SH, BG, BMK,VS

