## Supplementary material for "Variation in uptake and dose reduction of CDK4/6 inhibitors for the treatment of breast cancer in England, 2019–2024: a descriptive observational study using OpenPrescribing Hospitals"

**Table S1.** Treatments for invasive breast cancer that received positive appraisals for use in England by the National Institute for Health and Care Excellence between 2020 and 2024.

| Technology appraisal number | Technology appraisal title | Date of publication | Date last updated |
|---|---|---|---|
| TA952 | Talazoparib for treating HER2-negative advanced breast cancer with germline BRCA mutations | 21 February 2024 | 21 February 2024 |
| TA886 | Olaparib for adjuvant treatment of BRCA mutation-positive HER2-negative high-risk early breast cancer after chemotherapy | 10 May 2023 | 10 May 2023 |
| TA851 | Pembrolizumab for neoadjuvant and adjuvant treatment of triple-negative early or locally advanced breast cancer | 14 December 2022 | 14 December 2022 |
| TA836 | Palbociclib with fulvestrant for treating hormone receptor-positive, HER2-negative advanced breast cancer after endocrine therapy | 26 October 2022 | 26 October 2022 |
| TA819 | Sacituzumab govitecan for treating unresectable triple-negative advanced breast cancer after 2 or more therapies | 17 August 2022 | 17 August 2022 |
| TA816 | Alpelisib with fulvestrant for treating hormone receptor-positive, HER2-negative, PIK3CA-mutated advanced breast cancer | 10 August 2022 | 10 August 2022 |
| TA810 | Abemaciclib with endocrine therapy for adjuvant treatment of hormone receptor-positive, HER2-negative, node-positive early breast cancer at high risk of recurrence | 20 July 2022 | 20 July 2022 |
| TA801 | Pembrolizumab plus chemotherapy for untreated, triple-negative, locally recurrent unresectable or metastatic breast cancer | 29 June 2022 | 29 June 2022 |
| TA786 | Tucatinib with trastuzumab and capecitabine for treating HER2-positive advanced breast cancer after 2 or more anti-HER2 therapies | 27 April 2022 | 27 April 2022 |
| TA725 | Abemaciclib with fulvestrant for treating hormone receptor-positive, HER2-negative advanced breast cancer after endocrine therapy | 15 September 2021 | 15 September 2021 |
| TA687 | Ribociclib with fulvestrant for treating hormone receptor-positive, HER2-negative advanced breast cancer after endocrine therapy | 31 March 2021 | 31 March 2021 |
| TA639 | Atezolizumab with nab-paclitaxel for untreated PD-L1-positive, locally advanced or metastatic, triple-negative breast cancer | 1 July 2020 | 1 July 2020 |
| TA632 | Trastuzumab emtansine for adjuvant treatment of HER2-positive early breast cancer | 10 June 2020 | 10 June 2020 |

**Table S2.** Summary of licenses, NICE appraisals, and key trial outcomes for palbociclib, abemaciclib and ribociclib.

| Setting | Detail | | Palbociclib | Abemaciclib | Ribociclib |
|---|---|---|---|---|---|
| Advanced or metastatic | Licensed indication | | Treatment of HR+, HER2- locally advanced or metastatic breast cancer in combination with an aromatase inhibitor, or in combination with fulvestrant in women who have received prior endocrine therapy | Women with HR+, HER2- locally advanced or metastatic breast cancer in combination with an aromatase inhibitor or fulvestrant as initial endocrine-based therapy, or in women who have received prior endocrine therapy* | Women with HR+, HER2- locally advanced or metastatic breast cancer in combination with an aromatase inhibitor or fulvestrant as initial endocrine-based therapy, or in women who have received prior endocrine therapy |
| ‘First Line’ Metastatic | Licensing Trial | | PALOMA-2[1] | MONARCH-3[2] | MONALEESA-2[3] |
| | Treatment arms | | Palbociclib-letrozole vs placebo-letrozole | Abemaciclib-NSAI vs placebo-NSAI | Ribociclib-letrozole vs placebo-letrozole |
| | Outcome | PFS | 24.8m vs 14.5m (HR 0.58; 95% CI 0.46-0.72; P<0.001)[4] (Nov 2016) | Median not reached vs 14.7m (HR 0.54; 95% CI 0.41-0.72; P=0.000021)[5] (Oct 2017)<br><br>28.2m vs 14.7m (HR 0.54; 95% CI 0.42-0.70; P=0.000002)[6] (Jan 2019) | Median not reached vs 14.7m (HR 0.56; 95% CI 0.43-0.72; P = $3.29\times10^{-6}$ )[7] (Nov 2016)<br><br>25.3m vs 16.0m (0.57; 95% CI 0.46-0.70; log-rank P= 9.63 × 10-8)[8] (Jul 2018) |
| | | OS | 53.9m vs 51.2m (HR 0.96; 95% CI 0.78-1.18 ns)[9] (Jun 2022) | 66.8m vs 53.7m (HR 0.80; 95% CI 0.64-1.02; P= 0.0664 ns)[10] (May 2024)<br><br>63.7m vs 48.8m (HR 0.76; 95% CI 0.56-1.03; P=0.0757 ns)<br>Visceral subset | 63.9m vs 51.4m (HR 0.76; 95% CI 0.63-0.93; two-sided P=0.008)[11] (Mar 2022) |
| | NICE | TA | TA495[12] | TA563[13] | TA496[14] |
| | | Final appraisal | 16 November 2017 | 17 January 2019 | 16 November 2017 |
| | | Final guidance | 20 December 2017 | 27 February 2019 | 20 December 2017 |
| ‘Second Line’ Metastatic | Licensing Trial | | PALOMA-3[15] | MONARCH-2[16] | MONALEESA-3[17] |
| | Treatment arms | | Palbociclib-fulvestrant vs placebo-fulvestrant | Abemaciclib-fulvestrant vs placebo-fulvestrant | Ribociclib-fulvestrant vs placebo-fulvestrant |
| | Outcome | PFS | 9.5 m vs 4.6 m (HR 0.46; 95% CI 0.36-0.59; P < 0.0001)[18] (Mar 2016) | 16.4m v 9.3m (HR 0.553; 95% CI, 0.449-0.681; P < .001)[19] (Jun 2017) | 14.6m vs 9.1m (HR 0.57; 95% CI 0.43-0.74)[20] (Jun 2018) |
| | | OS | 34.9 m vs. 28.0 m (HR 0.81; 95% CI | 46.7m vs 37.3m (HR 0.76; 95% CI, | 39.7m vs 33.7m (HR 0.80; 95% CI |

| | | | | | |
|---|---|---|---|---|---|
| | | | 0.64-1.03; P = 0.09 ns)[21] (Oct 2018) | 0.61-0.95; P = .01)[22] (Sep 2019) | 0.61-1.05)[23] (Aug 2023) |
| | NICE | TA | TA619[24] (replaced by TA836[25]) | TA579[26] (replaced by TA725[27]) | TA687[28] |
| | | Final appraisal | 28 November 2019 | 02 April 2019 | 26 February 2021 |
| | | Final guidance | 15 January 2020 | 8 May 2019 | 31 March 2021 |
| Adjuvant | Licensed indication | | - | In combination with endocrine therapy is indicated for the adjuvant treatment of adult patients with HR+, HER2-, node+ early breast cancer at high risk of occurrence* | In combination with an aromatase inhibitor is indicated for the adjuvant treatment of patients with HR+, HER2- early breast cancer at high risk of occurrence** |
| | Licensing Trial | | PALLAS[29] | monarchE[30] | NATALEE[31] |
| | Treatment arms | | Palbociclib-ET vs ET alone | Abemaciclib-ET vs ET | Ribociclib-NSAI vs NSAI alone |
| | Outcome | IDFS | 4 year IDFS 84.2% vs. 84.5% (HR 0.96; 95% CI 0.81-1.14; P = 0.65 ns)[32] (Jan 2022) | 2 year IDFS 92.2% vs 88.7% (HR 0.75; 95%CI 0.60-0.93; P=0.01)[33] (Sep 2020)<br><br>5 year IDFS 83.6% vs 76.0% (HR 0.68; 95% CI 0.60-0.77); P<0.001)[34] (Mar 2024) | 3 year IDFS 90.4% vs 87.1% (HR 0.75; 95% CI 0.62–0.91; P = 0.003)[35] (Mar 2024) |
| | | OS | - | 86.8% vs 85.0% (HR 0.84; 95% CI 0.72-0.98); P 0.0273)[36] (Oct 2025) | - |
| | NICE | TA | - | TA810[37] | TA1086[38] |
| | | Final appraisal | - | 17 June 2022 | 24 April 2025 (node positive)<br>23 July 2025 (whole cohort) |
| | | Final guidance | - | 20 July 2022 | 06 August 2025 |

*In pre- or perimenopausal women, the endocrine therapy should be combined with a LHRH agonist.

**In pre- or perimenopausal women, or in men, the aromatase inhibitor should be combined with a LHRH agonist

HR+: Hormone Receptor positive
HER2: Human Epidermal Growth factor Receptor-2
LHRH: Luteinizing Hormone-Releasing Hormone
NSAI: Non-steroidal aromatase inhibitor
IDFS: Invasive disease-free survival

ns: non-sifnificant
PFS: Progression free survival
OS: Overall survival
NICE: National Institute for Health and Care Excellence
TA: Technology Appraisal

**Table S3.** Dosing schedule as specified in Summary of Product Characteristics, days treatment within 28 day cycle and WHO Defined Daily Dose for each CDK4/6 inhibitor.

| **CDK4/6 inhibitor** | | **Abemaciclib** | **Palbociclib** | **Ribociclib** |
|---|---|---|---|---|
| Dosing schedule | Initial starting dose | 150mg twice daily[39] | 125mg once daily[40*] | 600mg once daily[41] |
| | First dose reduction | 100mg twice daily | 100mg once daily | 400mg once daily |
| | Second dose reduction | 50mg twice daily | 75mg once daily | 200mg once daily |
| Days treatment within 28 day cycle | | Continuous | 21 days | 21 days |
| WHO Defined Daily Dose | | 300mg[42] (2x 150mg) | 94mg[43] (125mg x (21/28)) | 450mg[44] (600mg x (21/28)) |

* advanced or metastatic breast cancer.

**Table S4.** Total DDDs issued by NHS trusts within Cancer Alliances between January 2019 and December 2024.

| Cancer Alliance | Trust ODS Code | Trust Name | DDDs issued |
|---|---|---|---|
| Cheshire and Merseyside Cancer Alliance | REN | The Clatterbridge Cancer Centre NHS Foundation Trust | 617,373 |
| | RBT | Mid Cheshire Hospitals NHS Foundation Trust | 17,382 |
| | RJN | East Cheshire NHS Trust | 3,052 |
| | REM | Liverpool University Hospitals NHS Foundation Trust | 2,666 |
| East Midlands Cancer Alliance | RX1 | Nottingham University Hospitals NHS Trust | 315,112 |
| | RWE | University Hospitals Of Leicester NHS Trust | 170,719 |
| | RTG | University Hospitals Of Derby And Burton NHS Foundation Trust | 152,182 |
| | RWD | United Lincolnshire Teaching Hospitals NHS Trust | 119,290 |
| | RNS | Northampton General Hospital NHS Trust | 72,190 |
| | RNQ | Kettering General Hospital NHS Foundation Trust | 55,215 |
| | RFS | Chesterfield Royal Hospital NHS Foundation Trust | 689 |
| East of England Cancer Alliance | RDE | East Suffolk And North Essex NHS Foundation Trust | 273,649 |
| | RWH | East And North Hertfordshire Teaching NHS Trust | 256,526 |
| | RAJ | Mid And South Essex NHS Foundation Trust | 249,645 |
| | RC9 | Bedfordshire Hospitals NHS Foundation Trust | 108,775 |
| | RM1 | Norfolk And Norwich University Hospitals NHS Foundation Trust | 99,092 |
| | RGN | North West Anglia NHS Foundation Trust | 95,718 |
| | RGT | Cambridge University Hospitals NHS Foundation Trust | 80,434 |
| | RD8 | Milton Keynes University Hospital NHS Foundation Trust | 75,731 |
| | RQW | The Princess Alexandra Hospital NHS Trust | 66,422 |
| | RCX | The Queen Elizabeth Hospital, King's Lynn, NHS Foundation Trust | 57,907 |
| | RGR | West Suffolk NHS Foundation Trust | 49,447 |
| | RGP | James Paget University Hospitals NHS Foundation Trust | 25,745 |
| Greater Manchester Cancer Alliance | RBV | The Christie NHS Foundation Trust | 565,699 |
| | RRF | Wrightington, Wigan And Leigh NHS Foundation Trust | 59,374 |
| | RWJ | Stockport NHS Foundation Trust | 26,421 |
| | RMC | Bolton NHS Foundation Trust | 7,364 |
| Humber & North Yorkshire Cancer Alliance | RWA | Hull University Teaching Hospitals NHS Trust | 153,692 |
| | RCB | York And Scarborough Teaching Hospitals NHS Foundation Trust | 133,557 |
| | RJL | Northern Lincolnshire And Goole NHS Foundation Trust | 82,242 |
| Kent and Medway Cancer Alliance | RVV | East Kent Hospitals University NHS Foundation Trust | 173,365 |
| | RWF | Maidstone And Tunbridge Wells NHS Trust | 165,522 |
| | RPA | Medway NHS Foundation Trust | 73,972 |
| | RN7 | Dartford And Gravesham NHS Trust | 61,950 |
| Lancashire and South Cumbria Cancer Alliance | RXN | Lancashire Teaching Hospitals NHS Foundation Trust | 125,942 |
| | RXR | East Lancashire Hospitals NHS Trust | 121,203 |
| | RXL | Blackpool Teaching Hospitals NHS Foundation Trust | 112,166 |
| | RTX | University Hospitals Of Morecambe Bay NHS Foundation Trust | 101,786 |
| North Central London Cancer Alliance | RAL | Royal Free London NHS Foundation Trust | 187,674 |
| | RRV | University College London Hospitals NHS Foundation Trust | 76,942 |
| | RKE | Whittington Health NHS Trust | 26,419 |

| | | | |
|---|---|---|---|
| | RP4 | Great Ormond Street Hospital For Children NHS Foundation Trust | 131 |
| North East London Cancer Alliance | R1H | Barts Health NHS Trust | 209,368 |
| | RF4 | Barking, Havering And Redbridge University Hospitals NHS Trust | 105994.99 |
| Northern Cancer Alliance | RTD | The Newcastle Upon Tyne Hospitals NHS Foundation Trust | 134,315 |
| | RXP | County Durham And Darlington NHS Foundation Trust | 125,760 |
| | RTF | Northumbria Healthcare NHS Foundation Trust | 116,403 |
| | RTR | South Tees Hospitals NHS Foundation Trust | 95,678 |
| | RR7 | Gateshead Health NHS Foundation Trust | 91,814 |
| | R0B | South Tyneside And Sunderland NHS Foundation Trust | 86,277 |
| | RNN | North Cumbria Integrated Care NHS Foundation Trust | 85,029 |
| | RVW | North Tees And Hartlepool NHS Foundation Trust | 73,851 |
| Peninsula Cancer Alliance | REF | Royal Cornwall Hospitals NHS Trust | 174,897 |
| | RH8 | Royal Devon University Healthcare NHS Foundation Trust | 153,608 |
| | RK9 | University Hospitals Plymouth NHS Trust | 140,657 |
| | RA9 | Torbay And South Devon NHS Foundation Trust | 75,586 |
| Somerset, Wiltshire, Avon and Gloucestershire Cancer Alliance | RTE | Gloucestershire Hospitals NHS Foundation Trust | 204,004 |
| | RA7 | University Hospitals Bristol And Weston NHS Foundation Trust | 203,967 |
| | RH5 | Somerset NHS Foundation Trust | 141,644 |
| | RD1 | Royal United Hospitals Bath NHS Foundation Trust | 119,535 |
| | RNZ | Salisbury NHS Foundation Trust | 82,151 |
| South East London Cancer Alliance | RJ1 | Guy's And St Thomas' NHS Foundation Trust | 227,055 |
| | RJZ | King's College Hospital NHS Foundation Trust | 153,647 |
| | RJ2 | Lewisham And Greenwich NHS Trust | 60,706 |
| South Yorkshire and Bassetlaw Cancer Alliance | RHQ | Sheffield Teaching Hospitals NHS Foundation Trust | 329,459 |
| | RFF | Barnsley Hospital NHS Foundation Trust | 51,278 |
| | RFR | The Rotherham NHS Foundation Trust | 38,308 |
| | RP5 | Doncaster And Bassetlaw Teaching Hospitals NHS Foundation Trust | 903 |
| Surrey and Sussex Cancer Alliance | RA2 | Royal Surrey NHS Foundation Trust | 243,269 |
| | RYR | University Hospitals Sussex NHS Foundation Trust | 198,031 |
| | RXC | East Sussex Healthcare NHS Trust | 124,123 |
| | RTP | Surrey And Sussex Healthcare NHS Trust | 103,977 |
| | RDU | Frimley Health NHS Foundation Trust | 80,997 |
| | RTK | Ashford And St Peter's Hospitals NHS Foundation Trust | 159 |
| Thames Valley Cancer Alliance | RTH | Oxford University Hospitals NHS Foundation Trust | 195,113 |
| | RXQ | Buckinghamshire Healthcare NHS Trust | 96,507 |
| | RHW | Royal Berkshire NHS Foundation Trust | 95,571 |
| | RN3 | Great Western Hospitals NHS Foundation Trust | 87,772 |
| Wessex Cancer Alliance | RHU | Portsmouth Hospitals University NHS Trust | 170,655 |
| | R0D | University Hospitals Dorset NHS Foundation Trust | 140,978 |
| | RHM | University Hospital Southampton NHS Foundation Trust | 132,596 |
| | RN5 | Hampshire Hospitals NHS Foundation Trust | 116,480 |
| | RBD | Dorset County Hospital NHS Foundation Trust | 53,222 |
| | R1F | Isle Of Wight NHS Trust | 16,742 |
| West London Cancer Alliance | RPY | The Royal Marsden NHS Foundation Trust | 545,700 |
| | RYJ | Imperial College Healthcare NHS Trust | 170,086 |

| | | | |
|---|---|---|---|
| | RJ7 | St George's University Hospitals NHS Foundation Trust | 80,946 |
| | R1K | London North West University Healthcare NHS Trust | 60,003 |
| | RQM | Chelsea And Westminster Hospital NHS Foundation Trust | 39,907 |
| West Midlands Cancer Alliance | RRK | University Hospitals Birmingham NHS Foundation Trust | 338,647 |
| | RWP | Worcestershire Acute Hospitals NHS Trust | 172,968 |
| | RXW | The Shrewsbury And Telford Hospital NHS Trust | 154,732 |
| | RJE | University Hospitals Of North Midlands NHS Trust | 141,780 |
| | RL4 | The Royal Wolverhampton NHS Trust | 108,843 |
| | RKB | University Hospitals Coventry And Warwickshire NHS Trust | 104,412 |
| | RNA | The Dudley Group NHS Foundation Trust | 87,883 |
| | RLQ | Wye Valley NHS Trust | 76,387 |
| | RJC | South Warwickshire University NHS Foundation Trust | 56,380 |
| | RBK | Walsall Healthcare NHS Trust | 46,367 |
| | RLT | George Eliot Hospital NHS Trust | 42,986 |
| West Yorkshire and Harrogate Cancer Alliance | RR8 | Leeds Teaching Hospitals NHS Trust | 162,081 |
| | RWY | Calderdale And Huddersfield NHS Foundation Trust | 82,074 |
| | RXF | Mid Yorkshire Teaching NHS Trust | 81,781 |
| | RAE | Bradford Teaching Hospitals NHS Foundation Trust | 59,099 |
| | RCF | Airedale NHS Foundation Trust | 49,210 |
| | RCD | Harrogate And District NHS Foundation Trust | 48,344 |

**Table S5.** DDDs of each CDK4/6 inhibitor issued between January 2019 and December 2024 within trusts excluded from analysis.

| **Trust ODS Code** | **Trust Name** | **DDDs** | | |
|---|---|---|---|---|
| | | **Palbociclib** | **Abemaciclib** | **Ribociclib** |
| RTQ | Gloucestershire Health And Care NHS Foundation Trust | 0 | 28 | 0 |
| RV3 | Central And North West London NHS Foundation Trust | 0 | 149 | 123 |
| TAJ | Black Country Healthcare NHS Foundation Trust | 0 | -19* | 0 |

*Negative issuing quantities can occur due to *backtracking*. Not all stock that is issued within hospitals eventually ends up being used. Backtracking allows historical stock issues to be updated to reflect this.[45]

**Table S6.** Annual issuing quantity of individual CDK4/6 products across all included NHS trusts measured in (a) tablets/capsules (b) DDDs.

**(a)**

| Product | | 2019 | 2020 | 2021 | 2022 | 2023 | 2024 |
|---|---|---|---|---|---|---|---|
| Palbociclib (777047009) | Total | 740,305 | 976,727 | 1,292,959 | 1,427,294 | 1,341,196 | 1,270,227 |
| | 125mg capsules (33734211000001100) * | 445,014 | 528,179 | 76,095 | 12,153 | 6,960 | 3,741 |
| | 125mg tablets (39343811000001104) | 0 | 2,709 | 594,285 | 683,102 | 578,510 | 504,669 |
| | 100mg capsules (33734111000001108) * | 192,917 | 278,657 | 48,197 | 11,490 | 7,036 | 6,650 |
| | 100mg tablets (39343711000001104) | 987 | 3,990 | 345,551 | 425,798 | 420,349 | 402,246 |
| | 75mg capsules (33734311000001108) * | 101,387 | 163,171 | 2,8327 | 6,093 | 4,753 | 5,278 |
| | 75mg tablets (39343911000001104) | 0 | 21 | 200,504 | 288,658 | 323,588 | 347,644 |
| Abemaciclib (774399006) | Total | 321,504 | 595,325 | 636,442 | 912,826 | 2,316,109 | 3,967,588 |
| | 150mg tablets (36237111000001104) | 196,118 | 279,195 | 278,650 | 445,543 | 1,085,718 | 1,455,247 |
| | 100mg tablets (36237011000001104) | 85,050 | 221,440 | 251,748 | 325,140 | 890,601 | 1,668,758 |
| | 50mg tablets (36237211000001112) | 40,336 | 94,690 | 106,044 | 142,143 | 339,790 | 843,583 |
| Ribociclib (777435007) | Total | 239,007 | 375,835 | 389,107 | 548,535 | 976,565 | 1,450,469 |
| | 200mg tablets (34814111000001104) | 239,007 | 375,835 | 389,107 | 548,535 | 976,565 | 1,450,469 |

* Palbociclib capsules were phased out in January 2021

**(b)**

| Product | | 2019 | 2020 | 2021 | 2022 | 2023 | 2024 |
|---|---|---|---|---|---|---|---|
| Palbociclib (777047009) | Total | 878,949 | 1,136,863 | 1,492,922 | 1,624,915 | 1,495,190 | 1,392,659 |
| | 125mg capsules (33734211000001100) * | 591,774 | 702,366 | 101,190 | 16,161 | 9,255 | 4,975 |
| | 125mg tablets (39343811000001104) | 0 | 3,602 | 790,273 | 908,380 | 769,295 | 671,102 |
| | 100mg capsules (33734111000001108) * | 205,231 | 296,444 | 51,273 | 12,223 | 7,485 | 7,074 |
| | 100mg tablets (39343711000001104) | 1,050 | 4,245 | 367,607 | 452,977 | 447,180 | 427,921 |
| | 75mg capsules (33734311000001108) * | 80,894 | 130,190 | 22,601 | 4,861 | 3,792 | 4,211 |

| | | | | | | | |
|---|---|---|---|---|---|---|---|
| | 75mg tablets (39343911000001104) | 0 | 17 | 159,977 | 230,312 | 258,182 | 277,375 |
| Abemaciclib (774399006) | Total | 133,132 | 229,193 | 240,915 | 354,842 | 896,358 | 1,424,473 |
| | 150mg tablets (36237111000001104) | 98,059 | 139,598 | 139,325 | 222,772 | 542,859 | 727,624 |
| | 100mg tablets (36237011000001104) | 28,350 | 73,813 | 83,916 | 108,380 | 296,867 | 556,253 |
| | 50mg tablets (36237211000001112) | 6,723 | 15,782 | 17,674 | 23,691 | 56,632 | 140,597 |
| Ribociclib (777435007) | Total | 106,225 | 167,038 | 172,936 | 243,793 | 434,029 | 644,653 |
| | 200mg tablets (34814111000001104) | 106,225 | 167,038 | 172,936 | 243,793 | 434,029 | 644,653 |

* Palbociclib capsules were phased out in January 2021

**Table S7.** Percentage of each CDK4/6 inhibitor issued of all CDK4/6 inhibitors issued in 2021 and 2021, and the relative percentage change between these two periods across trusts within each Cancer Alliance in England.

| Cancer Alliance | Palbociclib | | | Abemaciclib | | | Ribociclib | | |
|---|---|---|---|---|---|---|---|---|---|
| | 2021 | 2024 | Change | 2021 | 2024 | Change | 2021 | 2024 | Change |
| Cheshire and Merseyside | 88.2 | 55.03 | -33.17 | 9.97 | 32.04 | 22.07 | 1.82 | 12.92 | 11.1 |
| Humber & North Yorkshire | 68.05 | 52.68 | -15.37 | 13.75 | 28.66 | 14.91 | 18.21 | 18.66 | 0.46 |
| Somerset, Wiltshire, Avon and Gloucestershire | 88.71 | 52.1 | -36.6 | 10.67 | 42.22 | 31.55 | 0.63 | 5.68 | 5.05 |
| Kent and Medway | 92.31 | 51.66 | -40.65 | 4.87 | 31.94 | 27.06 | 2.81 | 16.4 | 13.59 |
| North East London | 95.18 | 50.97 | -44.21 | 2.78 | 31.57 | 28.79 | 2.04 | 17.45 | 15.41 |
| West Yorkshire and Harrogate | 76.56 | 49.59 | -26.97 | 12.77 | 33.51 | 20.74 | 10.67 | 16.9 | 6.23 |
| Lancashire and South Cumbria | 79.23 | 42.74 | -36.48 | 16.14 | 48.09 | 31.95 | 4.63 | 9.17 | 4.54 |
| West Midlands | 75.35 | 40.49 | -34.86 | 18.42 | 44.09 | 25.67 | 6.22 | 15.42 | 9.19 |
| Northern | 84.98 | 39.99 | -44.98 | 12.2 | 36.33 | 24.13 | 2.83 | 23.68 | 20.85 |
| East of England | 76.02 | 39.95 | -36.08 | 9.36 | 37.34 | 27.98 | 14.62 | 22.72 | 8.1 |
| South Yorkshire and Bassetlaw | 86.46 | 39.44 | -47.02 | 6.68 | 38.37 | 31.68 | 6.85 | 22.19 | 15.34 |
| West London | 76.75 | 39.31 | -37.44 | 14.39 | 38.95 | 24.57 | 8.86 | 21.74 | 12.87 |
| Wessex | 71.27 | 37.39 | -33.88 | 19.23 | 48.46 | 29.23 | 9.49 | 14.15 | 4.65 |
| Peninsula | 76.22 | 35.79 | -40.43 | 7.69 | 30.97 | 23.29 | 16.1 | 33.24 | 17.14 |
| Thames Valley | 72.68 | 35.76 | -36.92 | 10.51 | 40.99 | 30.47 | 16.8 | 23.25 | 6.45 |
| Surrey and Sussex | 75.34 | 35.5 | -39.83 | 12.63 | 37.54 | 24.91 | 12.03 | 26.96 | 14.92 |
| East Midlands | 65.95 | 33.31 | -32.63 | 20.46 | 55.24 | 34.78 | 13.59 | 11.44 | -2.15 |
| North Central London | 84.92 | 28.93 | -55.99 | 6.34 | 48.27 | 41.93 | 8.74 | 22.8 | 14.06 |
| Greater Manchester | 79.42 | 25.8 | -53.62 | 13.39 | 57.65 | 44.26 | 7.19 | 16.54 | 9.35 |
| South East London | 65.01 | 25.43 | -39.58 | 15.82 | 45.95 | 30.13 | 19.17 | 28.62 | 9.45 |

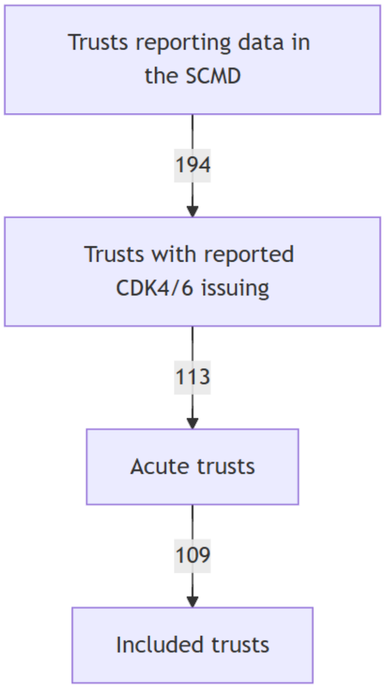


**Figure S1.** Flow diagram showing the number of NHS trusts included in the analysis.

39. Verzenios 150 mg film-coated tablets - Summary of Product Characteristics (SmPC). *electronic medicines compendium* https://www.medicines.org.uk/emc/product/11047/smpc.

40. IBRANCE 125 mg film-coated tablets - Summary of Product Characteristics (SmPC). *electronic medicines compendium* https://www.medicines.org.uk/emc/product/11962/smpc.

41. Kisqali 200 mg film-coated tablets - Summary of Product Characteristics (SmPC). *electronic medicines compendium* https://www.medicines.org.uk/emc/product/8110/smpc.

42. Abemaciclib - ATC/DDD Index. *WHO Collaborating Centre for Drug Statistics Methodology* https://atcddd.fhi.no/atc_ddd_index/?code=L01EF03.

43. Palbociclib - ATC/DDD Index. *WHO Collaborating Centre for Drug Statistics Methodology* https://atcddd.fhi.no/atc_ddd_index/?code=L01EF01.

44. Ribociclib - ATC/DDD Index. *WHO Collaborating Centre for Drug Statistics Methodology* https://atcddd.fhi.no/atc_ddd_index/?code=L01EF02.

45. Fisher, L., Black, S., MacKenna, B. & Speed, V. More about hospital stock control data. *Bennett Institute for Applied Data Science* https://www.bennett.ox.ac.uk/blog/2025/02/more-about-hospital-stock-control-data/ (2025) doi:10.53764/oph.oPoDVXRDFX.
